# No difference in cognitive task-related oscillations between human internal globus pallidus and subthalamic nucleus

**DOI:** 10.1101/2021.05.07.21256760

**Authors:** Muhammad Samran Navid, Stefan Kammermeier, Imran K. Niazi, Vibhash D. Sharma, Shawn M. Vuong, Jeremy D. W. Greenlee, Arun Singh

## Abstract

Recently it has been acknowledged that the basal ganglia nuclei play a major role in cognitive control; however, the contribution by their network remains unclear. Previous studies have demonstrated the role of the subthalamic nucleus (STN) in cognitive processing and suggested that its connections to cortical and other associated regions regulate response inhibition during conflict conditions. By contrast, the role of the internal globus pallidus (GPi) as the output nucleus before the thalamic relay has not yet been investigated during cognitive processing. We recorded local field potentials (LFPs) from externalized deep brain stimulation (DBS) electrodes implanted bilaterally in the GPi (n=9 participants with dystonia) and STN (n=8 participants with Parkinson’s disease (PD)) during a primed flanker task. Both dystonia (GPi group) and PD participants (STN group) responded faster to the congruent trials than the incongruent trials. Overall, the dystonic GPi group was significantly faster than the PD STN group. LFPs showed elevated cue-triggered theta (3-7 Hz) power in GPi and STN groups in a similar way. Response-triggered LFP beta power (13-25 Hz) was significantly increased in the GPi group compared to the STN group. Results demonstrate that GPi activity appears to be critical in the cognitive processing of action selection and response during the presence of conflict tasks similar to the STN group. As both GPi and STN nuclei are involved in cognitive processing; therefore, these nuclei may be targeted for neuromodulation to improve cognitive control via DBS.

## Introduction

The basal ganglia are a network of interconnected subcortical nuclei, which can play a critical role in cognitive processing. According to the classical basal ganglia network model, subthalamic nucleus (STN) activity suppresses the motor system by inhibiting the basal ganglia output structure, internal globus pallidus (GPi) by the indirect pathway (Alexander and Crutcher, 1990; Mink, 1996; Singh, 2018). As another gateway of the basal ganglia circuit, STN receives input from the frontal cortex via the hyperdirect pathway (Nambu et al., 2002). Through this hyperdirect pathway, cortical regions associated with executive functions directly influence both STN and GPi activity. Most recent studies have suggested that STN plays a crucial role in modulating responses during conflicting motor tasks by the basal ganglia circuitry (Brittain et al., 2012; Zavala et al., 2013). It has been hypothesized that STN and its connections to cortical regions and GPi can regulate response inhibition during conflict conditions (Brittain et al., 2012); however, the role of GPi in cognitive processing, such as conflict tasks, has not been investigated thoroughly.

Externalized deep brain stimulation (DBS) electrodes provide an exclusive opportunity to record the local field potential (LFP) activity of the human basal ganglia nuclei, including GPi and STN, the most commonly utilized target for DBS therapy in patients with dystonia and Parkinson’s disease (PD), respectively (Singh and Botzel, 2013; Singh et al., 2011a; Singh et al., 2011b; Singh et al., 2013). Previous studies have shown increased low-frequency oscillations in the delta (1-4 Hz) or theta (4-7 Hz) bands in the STN LFPs of PD participants at the onset of target cue during conflict situations (Brittain et al., 2012; Cavanagh et al., 2011; Kelley et al., 2018; Zavala et al., 2014). These studies suggest that cognitive tasks-related low-frequency oscillations, specifically in the theta-band, can propagate from frontal cortical regions such as the medial prefrontal cortex to STN via the hyperdirect pathway (Smith et al., 1998). Simultaneous recordings of single-neuron activities and LFPs in the STN revealed a modulation during cognitive processing in coherence with theta oscillations of the medial prefrontal cortex; it was suggested that theta frequency DBS might improve cognitive dysfunction in PD (Kelley et al., 2018). Previous scalp electroencephalogram (EEG) studies support this role of cue-triggered midfrontal theta activity during cognitive processing in PD participants (Singh et al., 2020; Singh et al., 2018). Together, these studies underline the significance of theta oscillations in the midfrontal cortical-STN network.

The glutamatergic STN projection neurons send their axons to the GPi and relay signals to the cerebral cortex through the thalamus in the indirect pathway. STN and GPi show motor- and cognitive tasks‐related activity in animal models and humans (Howell et al., 2016; Reymann et al., 2013) and affect frontal cortical activity (Devos et al., 2002; Yang et al., 2015). However, the differences between GPi and STN LFPs during cognitive processing are poorly understood. Here, we recorded LFP activity in the GPi and STN from dystonia and PD participants, respectively, who had been implanted with DBS electrodes. LFPs were collected via externalized DBS electrodes while participants performed a masked prime flanker task, which allowed us to understand the role of the GPi during conflict tasks and how it differs from STN activity. The masked prime flanker task is the higher-order conflict task because it envolves subliminal priming to manipulate action selection (Wenke et al., 2010).

## Materials and Methods

### Participants and surgery

A total of 17 participants (9 focal or regional dystonia and 8 PD) who had undergone bilateral DBS implantation in the GPi for dystonia and STN for PD participated in the study. STN participants were significantly older than the GPi participants (mean ± SD age: STN = 60 ± 4 years, GPi = 49 ± 13 years; independent t-test *p*=0.016). All clinical details are summarized in Table 1. All participants gave their written informed consent to participate in the study, which was approved by the local university’s ethics committee of the University Hospital Munich according to the Declaration of Helsinki.

**Table 1.** Clinical demographics.

| Subject | Gender | Disease | Analyzed electrode |  | Clinical score |  |
| --- | --- | --- | --- | --- | --- | --- |
|  |  | duration |  |  | PreOp. | PostOp. |
|  |  | (years) | L | R |  |  |
| GPi group (Dystonia participants), 30-63 years old |  |  |  |  | TWSTRS |  |
| 1 | M | 5 | 01 | 12 | 18 | 9 |
| 2 | M | 7 | 01 | 23 | 25 | 16 |
| 3 | F | 15 | 23 | 12 | 18 | 13 |
| 4 | F | 13 | 12 | 23 | 38 | 16 |
| 5 | F | 11 | 12 | 12 | 23 | 11 |
| 6 | M | 20 | 01 | 12 | 22 | 7 |
| 7 | M | 18 | 23 | 01 | 6 | 4 |
| 8 | M | 8 | 23 | 23 | 10 | 7 |
| 9 | M | 8 | 12 | 01 | 24 | 20 |
| STN group (Parkinson’s disease participants), 56-67 years old |  |  |  |  | Motor UPDRS |  |
| 1 | M | 12 | 23 | 12 | 25 | 9 |
| 2 | M | 8 | 12 | 12 | 26 | 10 |
| 3 | F | 9 | 23 | 12 | 28 | 12 |
| 4 | M | 8 | 12 | 01 | 40 | 29 |
| 5 | M | 10 | 01 | 23 | 29 | 16 |
| 6 | M | 8 | 12 | 01 | 14 | 10 |
| 7 | F | 20 | 23 | 12 | 36 | 24 |
| 8 | M | 11 | 01 | 23 | 28 | 15 |

The GPi-DBS electrodes used were type 3387 (Medtronic Inc, Minnesota, USA) with four platinum-iridium cylindrical connections (1.27 mm in diameter and 1.5 mm in length) and an inter-electrode separation of 1.5 mm. STN-DBS electrodes (model 3389, Medtronic Inc, Minnesota, USA) have a larger inter-contact separation of 0.5 mm. Contact 0 was designated the lowermost, and contact 3 was the uppermost in both types of DBS electrodes. The intended coordinates at the tip of contact 0 for GPi were 20 mm lateral from midline, 3 mm anterior to the mid-commissural point, and 3 mm below the anterior and posterior commissural (AC-PC) line based on the Talairach-Tournoux stereotactic atlas. The targeted coordinates for STN were 12 mm lateral, 3 mm posterior, and 4 mm below the midpoint of the AC-PC line. The intended coordinates of the GPi and STN were adjusted through direct visualization of the target nucleus on individual pre-operative stereotactic T2-weighted magnetic resonance imaging (MRI) and intraoperative microelectrode recordings. The final electrode position was confirmed in all participants by immediate postoperative MRI, which confirmed that at least one DBS electrode contact appeared within the target (Fig. 1A). Correct placement of the DBS electrode in the target nucleus was supported by improvement in Toronto Western Spasmodic Torticollis Rating Scale (TWSTRS) for dystonic patients and in motor Unified Parkinson’s Disease Rating Scale (UPDRS III) for Parkinson’s patients during ON DBS post-operatively compared to pre-operative status.

**Figure 1.**
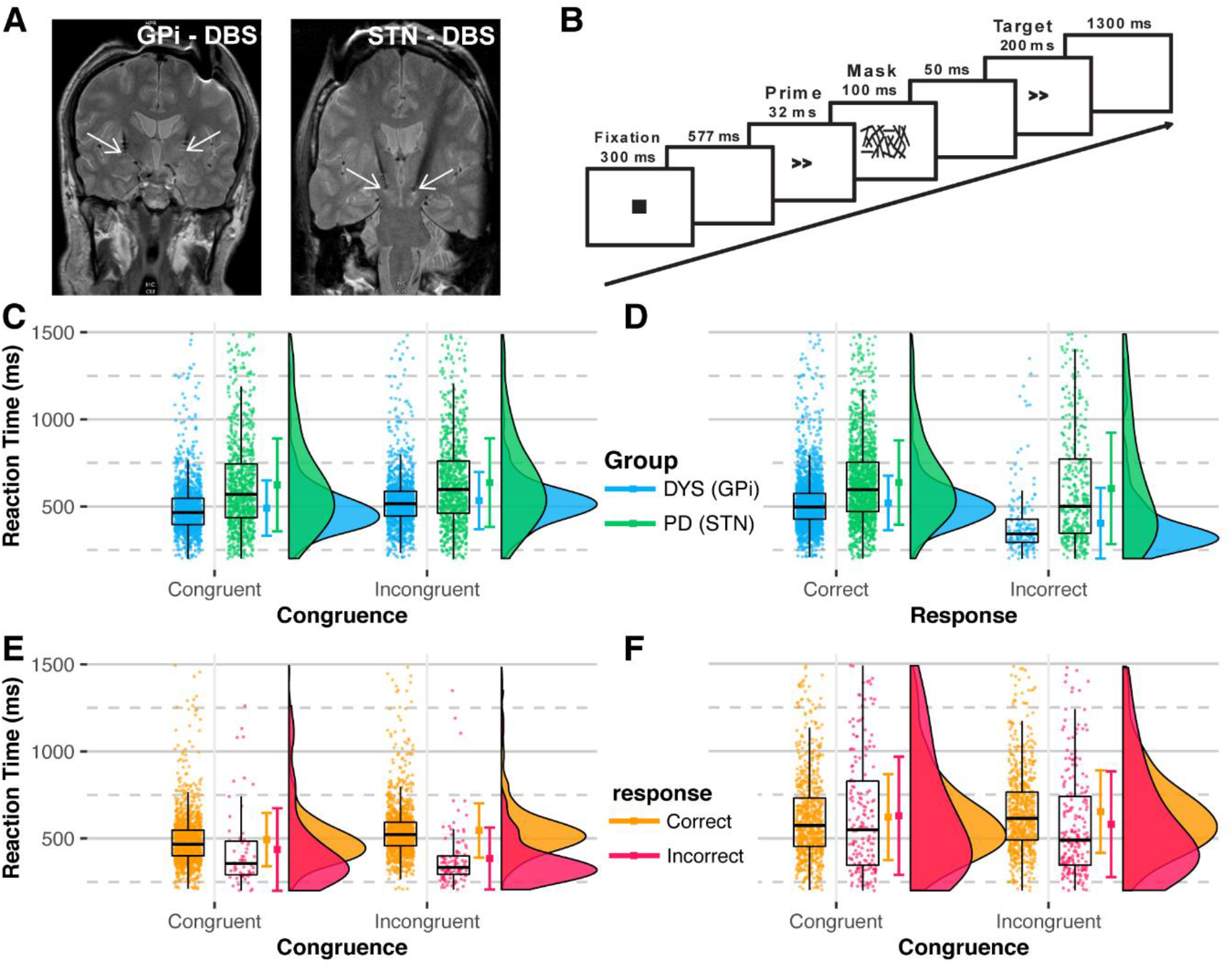
Location of DBS leads, experiment design, and reaction times (RTs). A) DBS electrodes were inserted in the GPi nuclei of dystonia and STN of PD participants for therapeutic purposes. White arrows show the lowermost contact of the DBS electrode. B) Trial design for the experiment. Prime and target arrows pointed in the same direction during the “congruent” trial and in the opposite direction during the “incongruent” trial. C) Graphs show reaction time RT displayed during congruent versus incongruent trials sorted by the participant group. There is a concentrated distribution of RT of the GPi group (dystonia participants) compared to the distribution of the STN group (PD participants) with more dispersed distribution, suggesting that the RTs of the GPi group were more consistent compared to the STN group. Additionally, the GPi group had shorter reaction times. D) Graphs display RT by the correctness of the response. As in (C), the GPi group had concentrated and quicker RTs than the STN group, irrespective of the correctness of the response. The RTs of the (E) GPi and (F) STN groups are shown, where RTs are similar for congruent and incongruent stimuli within each group; there were more consistent RTs for the GPi group compared to the STN group, which had widely distributed RTs. C-F: Dots represents RTs from all trials. The distribution plots show the density distribution estimated by a Gaussian kernel with SD of 1.5.

### Cognitive task and analysis

Figure 1B displays a schematic overview of the task structure. Each trial started with a fixation dot at the center of the screen for 300 ms. It was obscured ∼500 ms before the appearance of the prime stimulus. Prime stimuli were right or left-pointing double arrowheads (>> or <<), presented for 32 ms at the fixation point. Prime stimuli were followed by masking stimuli, consisting of random lines for 100 ms (Boy et al., 2010). Masking stimuli disappeared 50 ms before the appearance of the target stimulus. Target stimuli consisted of arrows similar to the prime stimuli in size and shape for the duration of 200 ms. The target stimulus conveyed the response instruction. The subsequent trial began 1300 ms after target offset. The stimuli were presented on a computer monitor in black on a white background. All stimuli were displayed at a visual angle of approximately 1° × 1°. Patients were seated in front of the computer screen ∼60 cm away from their eyes in the center of the horizontal line of sight. The experiment was executed in a dimly lit room. The experiment was comprised of six experimental blocks of 60 trials each. The task was preceded by a practice block of 20 trials. Each block contained the same number (30) of trials having a congruent and incongruent prime-target relationship and appeared in a pseudorandomized order. Patients were instructed to respond as fast and accurately as possible by pressing the arrow key with the hand corresponding to the direction of the target arrow. A response was considered correct if given within 200-1500 ms of the target cue (Duprez et al., 2019); otherwise, it was labeled as an incorrect response. Trials with no responses or responses outside this window were removed from further analysis.

### LFP recordings and analysis

All medication was stopped at least 10-12 hours in both dystonia and PD participants before the LFP recordings. LFPs were recorded from adjacent contact pairs of the DBS electrode (bipolar: 0-1, 1-2, and 2-3) using Brain Vision Recorder (Brain Products GmbH, Gilching, Germany) in the interval (2 days) between their DBS electrode implantation surgery and subsequent surgery for connection to the subcutaneous stimulator. LFP signals were sampled at 2.5 kHz and bandpass filtered from 0.1 Hz to 1 kHz.

All LFP data processing and analysis were performed in MATLAB R2018b (MathWorks). Left and right STN/GPi electrodes were assumed to be independent (Duprez et al., 2019; Zavala et al., 2013; Zavala et al., 2014); therefore, analyses were applied two times the number of subjects. Data from the active stimulation electrodes were analyzed. The data was down-sampled to 500 Hz and filtered between 1 to 50 Hz with 1812-ordered Kaiser windowed FIR filter (β = 5.65) at a transition bandwidth of 1 Hz. For identifying artifacts, baseline-corrected data (–500 to +3000 ms) with respect to the target stimulus was used, where baseline data was taken from a window of – 200 to –100 ms with respect to the prime stimulus. Any trial with absolute LFP amplitudes greater than the mean ± 4 SD was removed. For time-frequency analysis, data were epoched from –500 to +1500 ms with respect to the stimuli/response.

We computed the spectral measures by multiplying the fast Fourier transformed (FFT) power spectrum of each epoch with the FFT power spectrum of a set of complex Morlet wavelets (defined as a Gaussian-windowed complex sine wave: 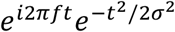, where *t* denotes time, and *f* is the frequency). The wavelets increased from 1 to 50 Hz in 50 logarithmically-spaced steps. These steps defined the width (or ‘cycles’) of each frequency band, increasing from three to ten cycles, between 1 and 50 Hz, and taking the inverse FFT (Singh et al., 2021; Singh et al., 2018). The result of this process was identical to signal convolution in the time domain and produced estimates of instantaneous power. The power was normalized by converting the spectral amplitude to a decibel (dB) scale using the formula 10 * *log*_10_(*power_t_/power_baseline_*), allowing us to compare the effects across frequency bands directly. As in our previous work, the baseline for each frequency was calculated by averaging power from −300 to −200 ms before the onset of the stimulus/response (Singh et al., 2020; Singh et al., 2021; Singh et al., 2018). Afterward, epochs were segmented from –500 to +1000 ms. The time-frequency region-of-interests (tf-ROIs) were selected a priori in the theta (3-7 Hz) and the beta (13-25 Hz) bands in the time window of 0 to +500 ms with respect to prime and target stimuli, and –100 to +400 ms with respect to response. These tf-ROIs were based on previous studies (Singh et al., 2020; Singh et al., 2021; Singh et al., 2018).

### Statistical analysis

The mixed-effect model was used to identify the effects of the disease, congruence, correctness of response, and their interactions on the reaction time (RT). The between-subject variance was estimated using random intercept in the model. The linear mixed model (using the lme function) and non-linear mixed model (using the glmer function) with gamma as link functions were tested. The models were implemented using lme4 version 1.1.23 (Bates et al., 2015) in R version 3.5.1. Since data and residuals were not normally distributed and had unequal variances, we used gamma distribution to model data. Additionally, a lower Akaike information criterion corrected (AICc) value favoured the use of a gamma-based model (AICc glmer = 57582.98; AICc lmer = 58893.08). The use of this model was supported by the research (Lo and Andrews, 2015) for analyzing RTs. The contrasts were obtained using the emmeans package adjusted for multiple comparisons using Tukey’s HSD. Two-way ANOVAs were used to identify any differences in the targeted tf-ROIs. Participant group (Dystonia, PD) and “congruence” (congruent, incongruent stimuli) or “response” (correct, incorrect) were used as factors. Pairwise comparisons were made using L SD correction. Statistical significance for all tests was set at 0.05.

## Results

### Behavioral results

The RTs from all trials in GPi and STN groups are shown in Figure 1C-F. The mixed model analysis showed significant interactions between groups, congruence, and correctness of response (Table 2). The RTs of the STN group were slower than those of the GPi group (Fig.1C-F, Fig. S1, and Table S1). By contrast, the difference between the RTs of correct and incorrect responses was larger for the GPi group. Pairwise comparisons showed a significant difference in RTs between the two groups irrespective of the congruence and correctness of response (all *p*<0.001) (Table S1). The response accuracy was 93.6% for the GPi group compared to 77.4% for the STN group.

**Table 2.** Mixed model results.

|  | Chisq | Df | p.value |
| --- | --- | --- | --- |
| participant | 134.11 | 1 | <0.001 |
| congruence | 119.32 | 1 | <0.001 |
| response | 411.95 | 1 | <0.001 |
| participant:congruence | 13.38 | 1 | <0.001 |
| participant:response | 82.21 | 1 | <0.001 |
| congruence:response | 407.82 | 1 | <0.001 |
| participant:congruence:response | 22.28 | 1 | < 0.001 |

### Cognitive task-related GPi and STN activities

Time-frequency analyses of the GPi LFPs and STN LFPs for prime cue-triggered, target cue-triggered congruent and incongruent trials, and response-triggered correct and error trials are presented in Figures 2, 3, and S2. Initially, we analyzed prime-triggered activity in the GPi and STN groups (Fig. S2) and compared between GPi/STN group and congruence for the two tf-ROIs aligned with respect to the prime. We observed a significant main effect at the group level in the theta-band (F(1,64)=6.65, *p*<0.05), but there was no significant difference between the GPi and STN activity due to congruence and interaction of participant group and congruence. ANOVA analysis showed no significant changes between the GPi and STN activity in the beta-band.

**Figure 2.**
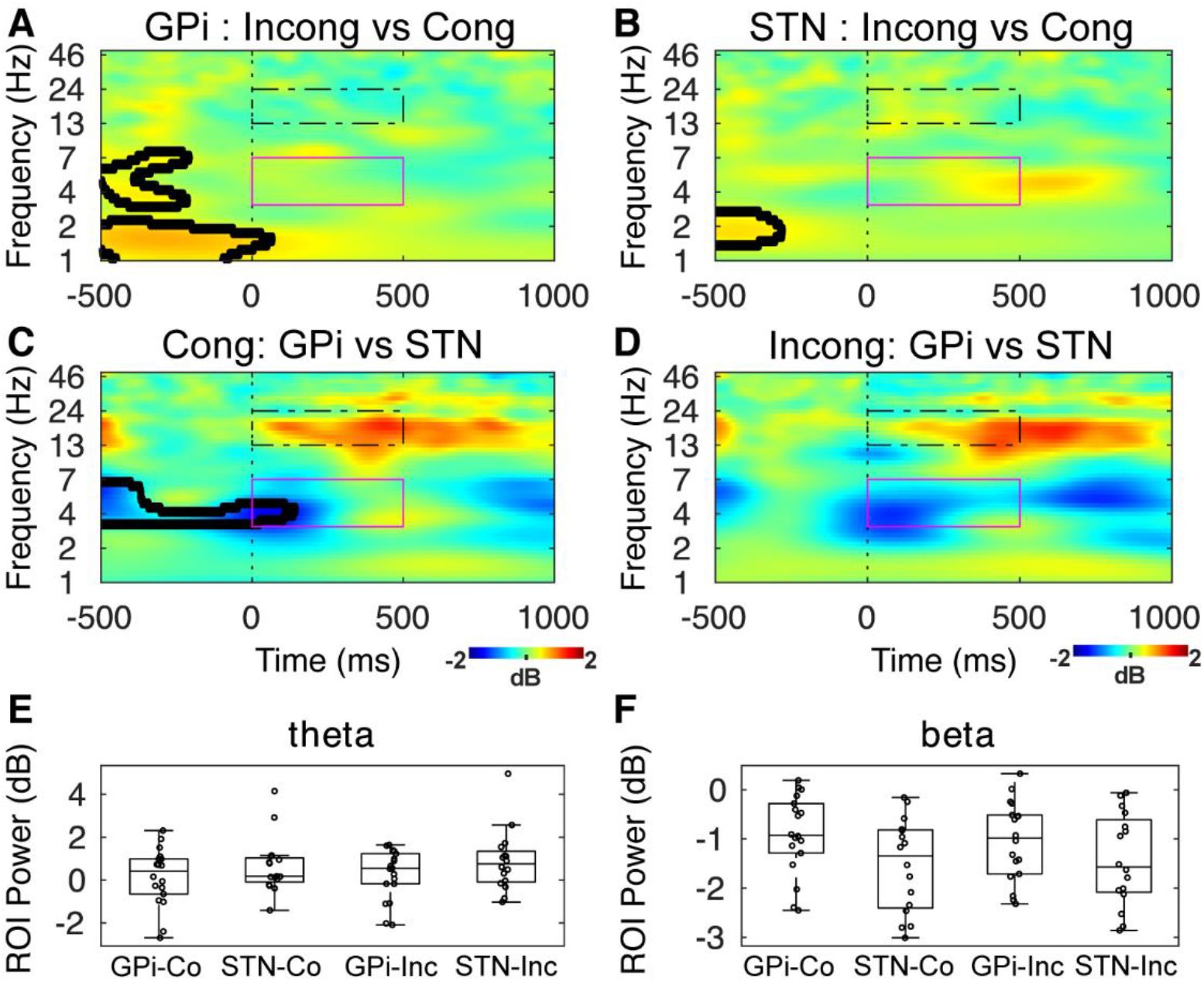
Target cue-triggered GPi and STN activity. No significant differences were seen between the GPi and STN activity in the tf-ROIs. The significant differences calculated by t-tests are enclosed in black lines. Magenta colored box = theta ROI; black dotted box = beta ROI. GPi: globus pallidus internus; STN: subthalamic nucleus; Co: Congruent; Inc: Incongruent.

**Figure 3.**
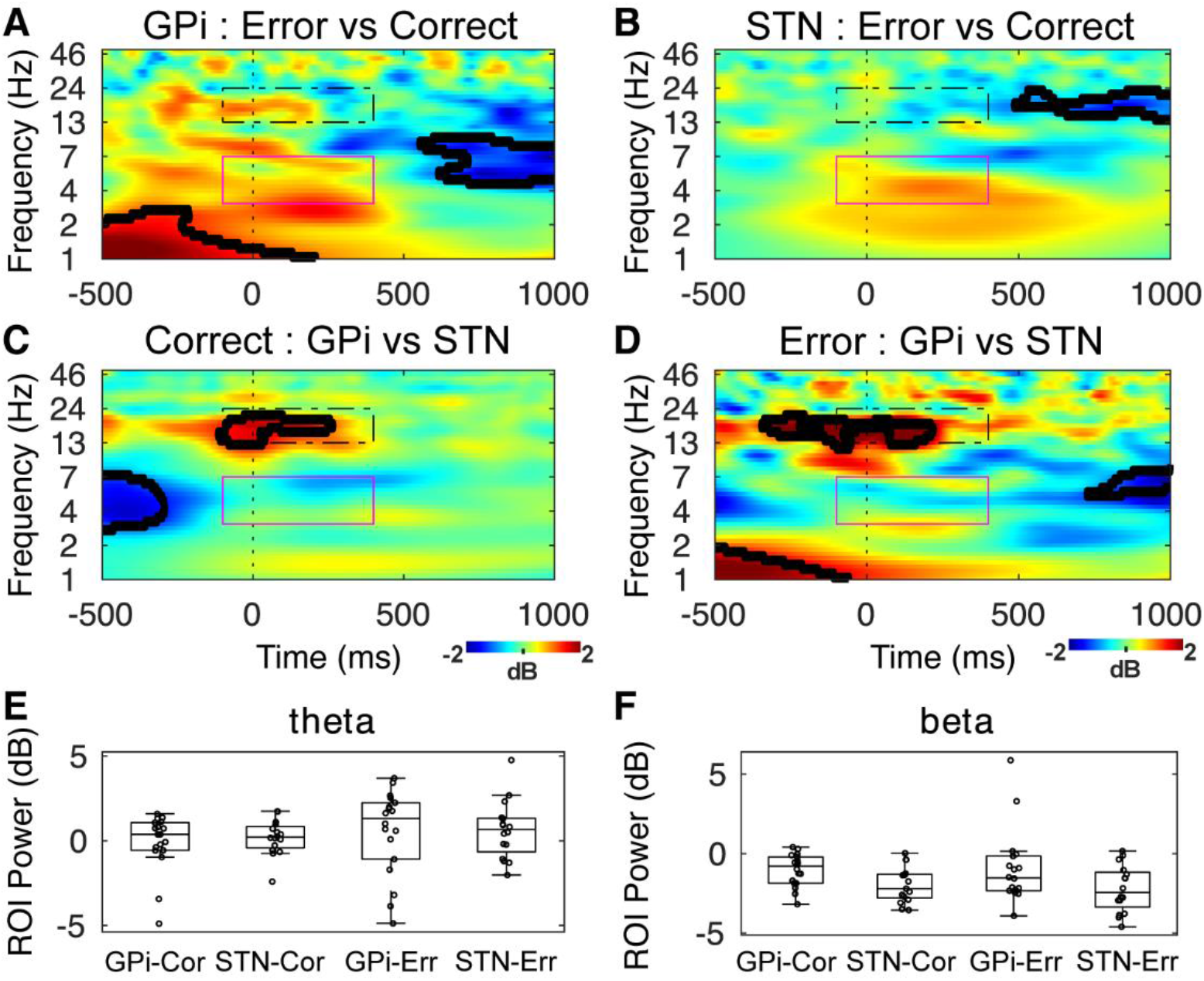
Response-triggered GPi and STN activity. Significant differences were seen between the GPi and STN activity in the beta ROI. The significant differences calculated by t-tests are enclosed in black lines. Magenta colored box = theta ROI; black dotted box = beta ROI. GPi: globus pallidus internus; STN: subthalamic nucleus; Co r: Correct; Err: Error.

Next, we analyzed target cue-triggered evoked activities in the time-frequency plots (Fig. 2) and compared between GPi/STN groups and congruence for the two ROIs aligned with respect to the cue. We observed no differences in the power of GPi and STN activity in the theta ROI when data was aligned with respect to the target cue. However, there were differences in the GPi and STN activity at the group level in the beta ROI (main effect: F(1,64)=4.39, *p* < 0.05). There was no effect of congruence and interaction of group and congruence on the power in the beta ROI. A previous study has shown the increased activity in the theta-band power in the STN group around 500 ms after the target cue when incongruent trials were compared with congruent trials (Zavala et al., 2013). Our data did not only show a similar oscillatory pattern in the STN group but also demonstrated that there is no difference in the theta-band activity between the STN and GPi group, suggesting the occurrence of cognitive task-related oscillatory patterns in the STN and GPi regions by the subthalamic-pallidal (STN-GPi) network (Magill et al., 2000).

Additionally, we analyzed response-triggered GPi and STN activities (Fig. 3) and compared between the GPi and STN groups and congruence for the two respective ROIs aligned with respect to the response or execution of the cognitive task. There was a significant difference in the group (main effect: F(1,64)=8.51, *p* = 0.005) based on the beta ROI, but not in the theta ROI. No congruence and interaction effects of group and congruence on the GPi and STN activity could be observed. Similar to our results, previous studies demonstrated response-locked desynchronization in the beta-band in the STN group (Brittain et al., 2012; Kuhn et al., 2004; Singh, 2018). Our prior studies have also demonstrated movement-related beta power changes in the GPi group (Singh and Botzel, 2013; Singh et al., 2011a).

## Discussion

Our results suggest the similarity between GPi and STN LFP activity at the onset of a target cue during cognitive processing since both nuclei displayed a similar pattern in the theta-band during the primed flanker task. Behavior results indicate that the GPi group performed the cognitive task with a higher speed and fewer errors compared to the STN group. Furthermore, the response-triggered LFP beta power was higher in the GPi group compared to the STN group.

To our knowledge, LFP recordings from the GPi and STN regions have not been reported during the primed flanker cognitive task. We designed our study not to observe disease-specific LFPs from the target region but instead to study the GPi LFPs during the cognitive task in comparison to STN LFPs. As one of the limitations of the current study, we cannot eliminate the disease factor from both groups; this could influence the oscillatory nature of both nuclei. Well-known abnormal oscillations among the STN datasets could be observed in the beta-band (George et al., 2013; Giannicola et al., 2010; Kühn et al., 2008), and in the alpha-band (8-12 Hz) among the GPi data (Singh et al., 2011b; Weinberger et al., 2012). The purview of this study focused on the theta-band during cognitive processing. We suggest that a similar kind of modulation in the theta-band oscillations in the GPi and STN during primed flanker task likely reflects cognitive processing via cortico-basal ganglia circuits (Jahanshahi et al., 2015; Magill et al., 2000). Previous EEG studies have reinforced the role of midfrontal theta oscillations as the mechanism of cognitive control and attenuated midfrontal theta power; this effect was seen in PD patients in comparison to healthy control subjects (Singh et al., 2021; Singh et al., 2018). STN LFPs have also shown this modulation in the theta-band during flanker and interval-timing cognitive tasks in PD patients (Lam et al., 2021; Zavala et al., 2013). The human STN is monosynaptically connected with cognitive brain areas such as the prefrontal cortex. A previous study has demonstrated that STN single neurons and LFPs are modulated during cognitive processing in coherence with theta oscillations in the medial prefrontal cortex (Kelley et al., 2018), suggesting an essential role of theta oscillations in the fronto-basal ganglia pathway during cognitive processing.

Frontal cortical, STN, and GPi regions as part of the fronto-striato-subthalamic-pallidal networks have been revealed to contribute to goal-directed and habitual inhibition (Jahanshahi et al., 2015). Subthalamic-pallidal interactions appear to be critical differentiators between normal and abnormal cognitive and motor functioning of the basal ganglia (Gillies et al., 2002). Connections between the STN and GPi are excitatory, and there is strong evidence that STN neurons strongly affect neuronal activity in the GPi (Nambu et al., 2000). In concordance with our current cognitive study, a previous study of our group (Singh et al., 2011b) has also shown the parallel STN and GPi oscillations in the alpha-band during motor performance. Therefore, the results suggest that not only STN but also GPi could be a promising therapeutic target to modulate oscillatory activity in the fronto-striato-subthalamic-pallidal networks with the intent to improve cognitive dysfunction in patients.

Our findings concerning theta-band activity suggest its role in cognitive mechanisms, and these are likely dopamine-independent since levodopa therapy does not improve theta power and cognitive behavior in PD participants (Singh et al., 2021; Singh et al., 2018). In accordance with current STN LFP findings, a prior report has revealed a novel role for the theta phase at the time of target cue during conflict monitoring and demonstrated a negative correlation between theta phase alignment and reaction time, reinforcing the role of theta-oscillation occurrence during conflicting tasks (Zavala et al., 2013). In GPi LFPs, on the other hand, theta oscillations have been related to the detection and assessment of errors in the flanker task (Herrojo Ruiz et al., 2014). Moreover, theta-band spectral power increased in GPi during erroneous responses, in line with the more significant negative GPi event-related potential component that emerged ipsilateral to overt errors. This error signal precedes the occurrence of theta power, suggesting that the GPi as the main basal ganglia output may contribute to the processing of motor error evaluation by the midfrontal region (Cavanagh et al., 2009; Singh et al., 2018).

An additional observation was b eta-band activity in the basal ganglia during the cognitive task-related motor responses. Our results revealed a decreased beta-band power at the time of both correct and erroneous responses in the STN group compared to the GPi group. A previous study has also demonstrated stimulus-triggered decreased STN beta-band activity lasting throughout the verbal response in the Stroop task, in alignment with the theory that previously elevated beta-band activity needs to decrease in order to allow a specific motor output to occur at the time of response (Brittain et al., 2012). More specifically, increased STN beta-band activity has been seen during “NoGo” trials and also in the associated frontal cortical regions during successful stopping in the stop-signal task (Kuhn et al., 2004; Swann et al., 2009). The GPi LFP in dystonia patients showed a reward-related response in the high beta/low gamma range (Munte et al., 2017). Our previous studies have also reported motor-response-related modulation in the beta-band in the GPi group (Singh and Botzel, 2013; Singh et al., 2011a). Altogether, these studies affirmed a close relationship between frontal cortical-STN-GPi networks and the beta-band activity during the motor responses. Our data may be seen in support of the concept connecting GPi and STN as substantial contributors to the application of cognitive tasks. These nuclei might potentially be targeted by modulatory DBS therapy to improve those functions in patients.

In the current study, it has to be noted that the STN group involved PD patients with bradykinesia, i.e., slowness of movements (Moustafa et al., 2016) compared to the GPi group that included dystonia patients who do not show bradykinetic features (Foley et al., 2017). In the advanced stage of PD, patients exhibit higher impairment in cognitive function (Aarsland et al., 2017; Singh et al., 2018); however, in the current study, participants with moderate to severe cognitive deficits were excluded in order to compare basal ganglia oscillatory activity between STN and GPi groups in cognitive processing. Similar to previous studies, our GPi group or dystonia patients did not exhibit any cognitive impairments or deficits in executive function or working memory, as confirmed by our expert movement disorder at the time of recruitment (Jahanshahi et al., 2003; Kuyper et al., 2011). Overall, both GPi and STN groups did not show any clinical neuropsychological impairments. Here, behavioral differences in cognitive performance between groups were observed likely due to the age of the participants. In the current study, the STN group was significantly older than the GPi group. The influence of cognitive performance in our STN group may be sufficiently explained by an age-dependent effect, as demonstrated elsewhere (Brigham and Pressley, 1988; Scheibe and Blanchard-Fields, 2009).

We acknowledge other limitations in this work. Previous studies in dystonia and PD patients were primarily focused on motor function, and fewer have studied the cognitive function of GPi and STN. We performed our experiments to gain insight into the normal cognitive function of the GPi and STN structures. All human studies are restricted to collect deep brain LFPs from groups of patients with severe neurological diseases during medically indicated procedures instead of normal healthy participants. In contrast, comparative animal studies have to rely on mostly insufficient comparable model diseases (e.g., MPTP-induced Parkinsonism), on top of distinct neuroanatomical and biological differences. Our dataset is exceptional as, in many cases, DBS electrodes are not commonly externalized. In current practice, many participants are operated on within a day to implant the impulse generator. This precludes a larger sample size data collection, which would be desirable. Regrettably, we could not investigate the effects of DBS on LFPs and cognitive behavior since it was not yet technically feasible to stimulate and record LFPs from the same active lead. However, in the future, the Percept™ PC device with the BrainSense technology (Medtronic, Minneapolis, USA) may be used to capture LFPs using the implanted DBS electrode. This will likely help in collecting LFPs from large numbers of DBS patients during clinically effective stimulation in cognitive performances.

## Data Availability

Data will be available upon a reasonable request.

## Acknowledgments

We thank Prof. Kai Boetzel from the Ludwig Maximilian University of Munich, Germany, for his guidance in collecting data and encouraging patients to participate in the research project. This work is supported by the Division of Basic Biomedical Sciences and Center for Brain and Behavior Research at the University of South Dakota, Vermillion, SD, USA.

## Author Contributions

Study design and concept: M.S.N., S.K., I.K.N., V.D.S., S.M.V., J.D.W.G., A.S; data collection: S.K. A.S.; data analysis: M.S.N., S.K., I.K.N., A.S.; drafting and revision of the manuscript: M.S.N., S.K., I.K.N., V.D.S., S. N.V., J.D.W.G., A.S.

## Data Availability

The data and analysis codes will be available from the corresponding author upon reasonable request.

## Supplementary Data

Additional supplementary information can be found in the online version of this article at the publisher’s website.

## Supplementary Information

### Supplementary Figures

**Figure S1.**
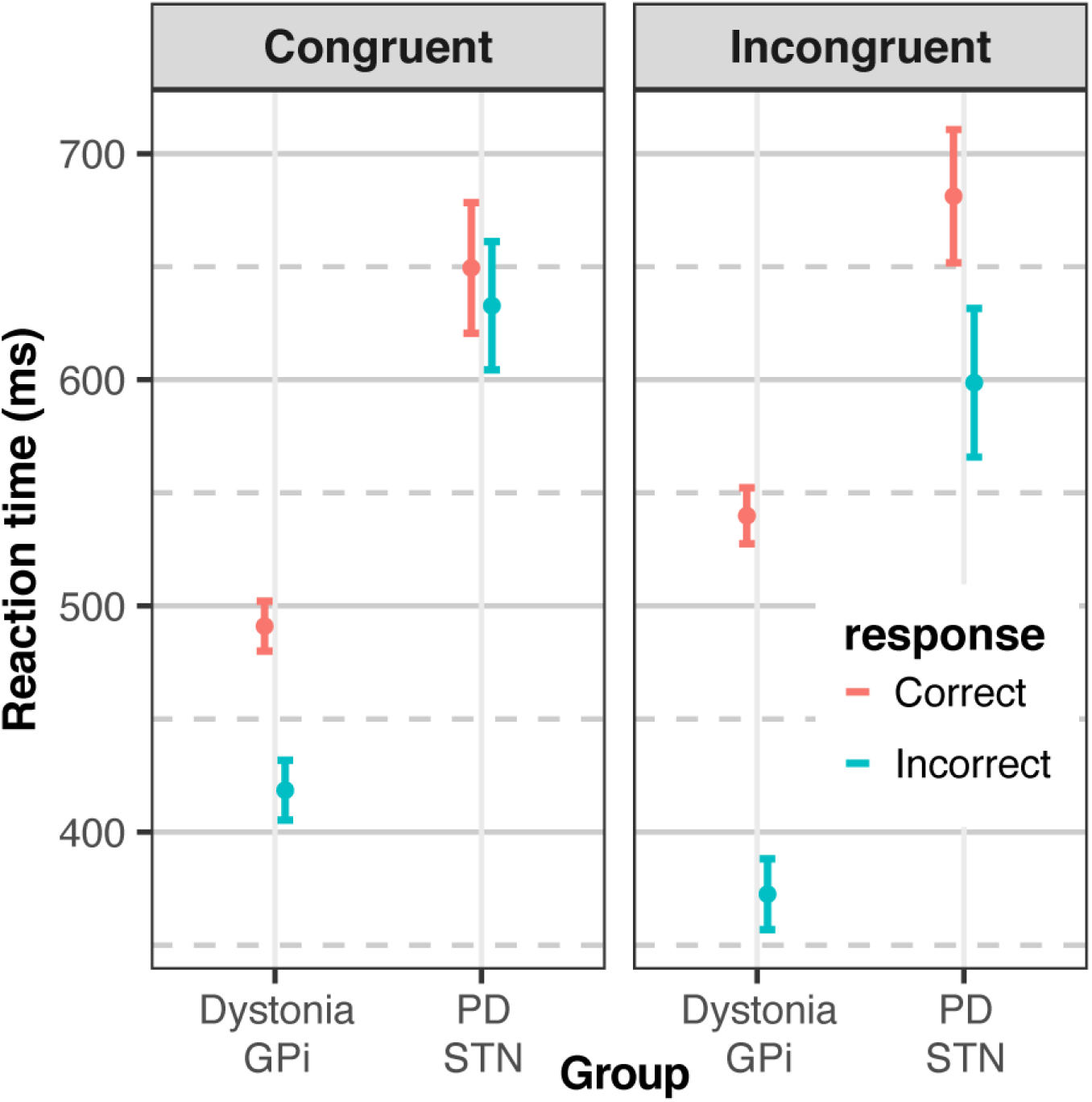
Estimated reaction times. The error bars represent mean ± 95% CI. GPi: globus pallidus internus; STN: subthalamic nucleus;

**Figure S2.**
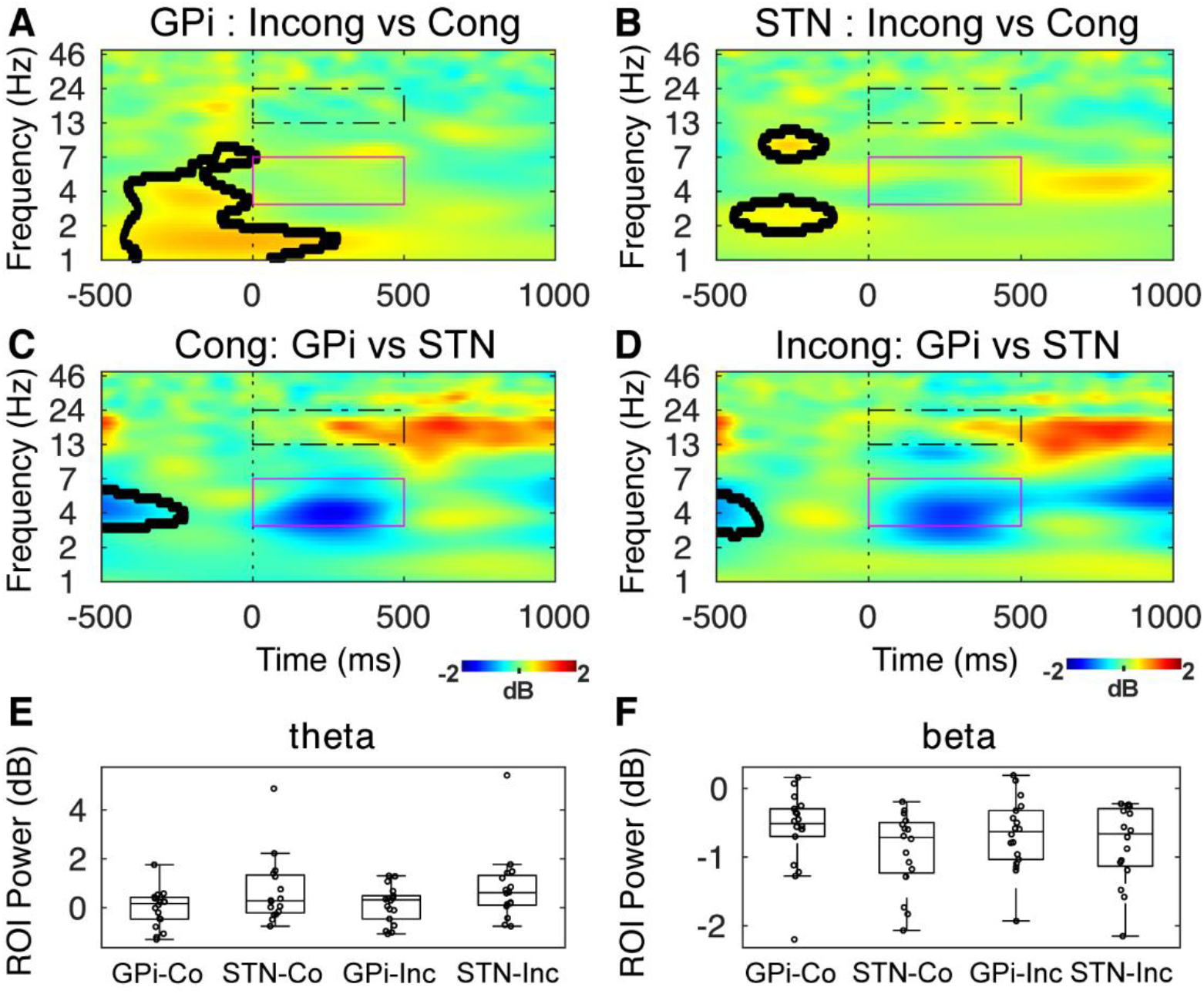
Prime-triggered GPi and STN activity. No significant differences were seen between the GPi and STN activity in the prime-triggered tf-ROIs. The significant differences calculated by t-tests are enclosed in black lines. Magenta colored box = theta ROI; black dotted box = beta ROI. GPi: globus pallidus internus; STN: subthalamic nucleus; Co: Congruent; Inc: Incongruent.

### Supplementary Table

**Table S1.** Differences between participant groups based on estimated reaction times (ms) from the statistical model.

| contrast | congruence | response | Estimate ± SE | LCL | UCL | z.ratio | p.value |
| --- | --- | --- | --- | --- | --- | --- | --- |
| GPI - STN | Congruent | Correct | -158.44 ± 13.86 | -185.60 | -131.27 | -11.43 | <0.001 |
| GPI - STN | Incongruent | Correct | -141.29 ± 14.02 | -168.77 | -113.81 | -10.08 | <0.001 |
| GPI - STN | Congruent | Incorrect | -214.25 ± 14.62 | -242.91 | -185.58 | -14.65 | <0.001 |
| GPI - STN | Incongruent | Incorrect | -226.14 ± 16.26 | -258.01 | -194.27 | -13.91 | <0.001 |

